# Privacy Protection for Chinese Electronic Medical Records Using Large Language Models: Effectiveness Evaluation and Application of LLM Models in Medical Data Tasks

**DOI:** 10.1101/2025.07.27.25332177

**Authors:** Gong Mengchun, Ouyang Zihao, Ma Dandan, Cai Endi, Liu Chao, Shi Wenzhao, Zhang Bohan, Ma Lian, Wei Yuna, Jiang Huizhen, Zhou Xiang

**Author notes:** Corresponding Author Zhou Xiang. These authors equally contributed to this study (Gong Mengchun, Ouyang Zihao, Ma Dandan).

## Abstract

**Background:** The privacy protection of medical patients has remained a critical concern in healthcare information management during the digital era. Conventional approaches have predominantly relied on rule-based protocols and data encryption systems, which typically require substantial involvement of IT professionals for implementation. Recent advancements in Large Language Models (LLMs) have introduced novel approaches for electronic medical records (EMRs) privacy protection, simultaneously enabling clinical practitioners to utilize these tools for specific data tasks.

**Objectives:** This study aims to leverage LLMs through a no-code framework to achieve structured processing of patient privacy data in Chinese EMRs and formulate privacy policies, while evaluating the practical efficacy of LLMs.

**Methods:** This study employs a disease-specific data subset from Peking Union Medical College Hospital (PUMCH), comprising data from approximately 160,000 patients, using a prompt engineering approach to enable LLMs to perform sensitive information annotation in lengthy EMR narratives. Simultaneously, it automates the classification of privacy-level for identified sensitive data and develops targeted protection strategies based on risk tiers, thereby mitigating non-essential exposure of patient privacy during data sharing. The research utilizes the Qwen model, with its entire workflow being exclusively driven by medical natural language prompts and self-evolving knowledge bases, requiring no supplementary programming or code development. These strategies were validated using the hospital’s test text dataset, with primary evaluation metrics focusing on precision rates (including accuracy of information extraction and privacy-level classification) and recall rate assessments for critical sensitive data categories.

**Results:** Utilizing 4 million text entries from PUMCH, we conducted sampled data observation and performed privacy annotation via LLM prompts across seven categories: names, addresses, contact details, national ID numbers, hospital names, sexually transmitted disease (STD) information, and pregnancy-related patient data. Through iterative prompt refinement via error analysis, optimal performance was achieved on the test set, demonstrating an average precision of 97% and recall of 95% across these seven entity types. Furthermore, sensitivity tier classification was implemented for three high-risk categories: addresses, STD information, and pregnancy-related data, attaining average precision of 95% and recall of 90% in sensitivity-level determination.

**Discussion:** We propose a novel codeless privacy protection framework leveraging LLMs, enabling intelligent anonymization of medical data through natural language interaction. This solution employs a three-tiered hierarchical protection mechanism that dynamically adapts privacy strategies to clinical scenario requirements, ensuring data security while maximizing data utility.

## Introduction

The evolution of electronic medical records (EMRs) has transformed how healthcare institutions store, access, and share patient data. Modern EMR systems integrate clinical information in digitized, standardized formats, enhancing data integrity and usability [1]. Concurrently, this advancement enables cross-institutional data sharing, which has become a critical driver for leveraging real-world evidence to complement clinical trial outcomes and healthcare capabilities across institutions, regions, and even nations [2]. Multi-center data utilization accelerates insights into disease patterns and treatment efficacy while facilitating longitudinal analyses of patient trends across demographics, thereby advancing epidemiological research, risk prediction, and personalized care[3].Advancing precision medicine and population health research.

However, patient privacy breaches during data sharing remain a pivotal challenge. Inadequate protection of personally identifiable information (PII) and sensitive health data—including diagnoses and treatment for critical conditions—can severely impact patients. Unauthorized access may lead to social stigmatization, employment or insurance discrimination, and significant emotional distress. Furthermore, such breaches erode public trust in healthcare systems, deterring patients from sharing crucial data in the future, ultimately jeopardize both individual well-being care and public health initiatives[4,5].

Data anonymization—a process involving the modification or “masking” of sensitive fields to reduce identifiability while preserving clinical utility—represents an underexplored yet promising approach. While many studies focus on heavy-duty techniques like encryption, access control, federated learning, and differential privacy, anonymization enables finer-grained control over what patient information is shared and how. By precisely targeting specific data types and scopes, healthcare organizations can maximize data sharing for research and operational purposes while minimizing privacy risks [6].

Traditional anonymization typically relies on rudimentary models that first identify sensitive entities and then mask or transform them. In practice, this often involves entity recognition using regular expression-based methods or basic machine learning techniques applied to patient data. Once extracted, sensitive entities undergo systematic anonymization to mitigate re-identification risks while retaining analytical utility. These conventional approaches remain popular due to their simplicity, though they may lack the contextual nuance captured by advanced methods. Nevertheless, they remain essential components of many data-sharing workflows for ensuring patient privacy [7,8].

Our study leverages large language models (LLMs) to advance data anonymization. LLMs offer a transformative alternative to traditional methods. Unlike rule-based regex or conventional machine learning models, LLMs harness advanced transformer architectures to better capture contextual subtleties in clinical narratives [9]. This enhanced contextual understanding improves identification accuracy for sensitive entities embedded in unstructured text, such as patient names, birthdates, and other identifiers. Crucially, through natural language prompting, clinical researchers can directly guide the anonymization process. This user-friendly approach eliminates dependency on technical experts, democratizes access to sophisticated anonymization tools, and enables more efficient, accurate patient data sharing without compromising privacy [10].

The application of LLMs in medicine marks a significant advancement, particularly in empowering clinicians to perform natural language processing (NLP) tasks—such as data anonymization, information extraction, and summarization—that previously required specialized expertise. By interacting with LLMs via natural language prompts, clinicians can now execute these tasks, making advanced NLP capabilities widely accessible.

Our goal is to utilize LLMs combined with prompt engineering and Retrieval-Augmented Generation (RAG) to achieve structured extraction of patient privacy data from Chinese EMRs and formulate corresponding protection strategies, while systematically evaluating model performance throughout this process.

## Methods

### Data Source

This study is based on a multicenter clinical research infrastructure developed through PUMCH’s disease-specific data platform (Figure 1). The system rigorously adheres to the Observational Medical Outcomes Partnership (OMOP) Common Data Model (CDM) standard, integrating comprehensive, multidimensional clinical data from 160,000 patients with selected diseases collected between 2012 and 2025. At the data architecture level, structured data undergo standardized mapping via OMOP-CDM specifications, while unstructured textual data are uniformly stored in a NOTE table containing 13 categories of core clinical documentation: outpatient/emergency records, ward round notes, discharge summaries, admission notes, initial progress note, 24-hour admission/discharge records, preoperative discussion notes, surgical reports, initial postoperative progress note, specialty-specific notes, periodic summaries, resuscitation records, and death certificates.

**Figure 1.**
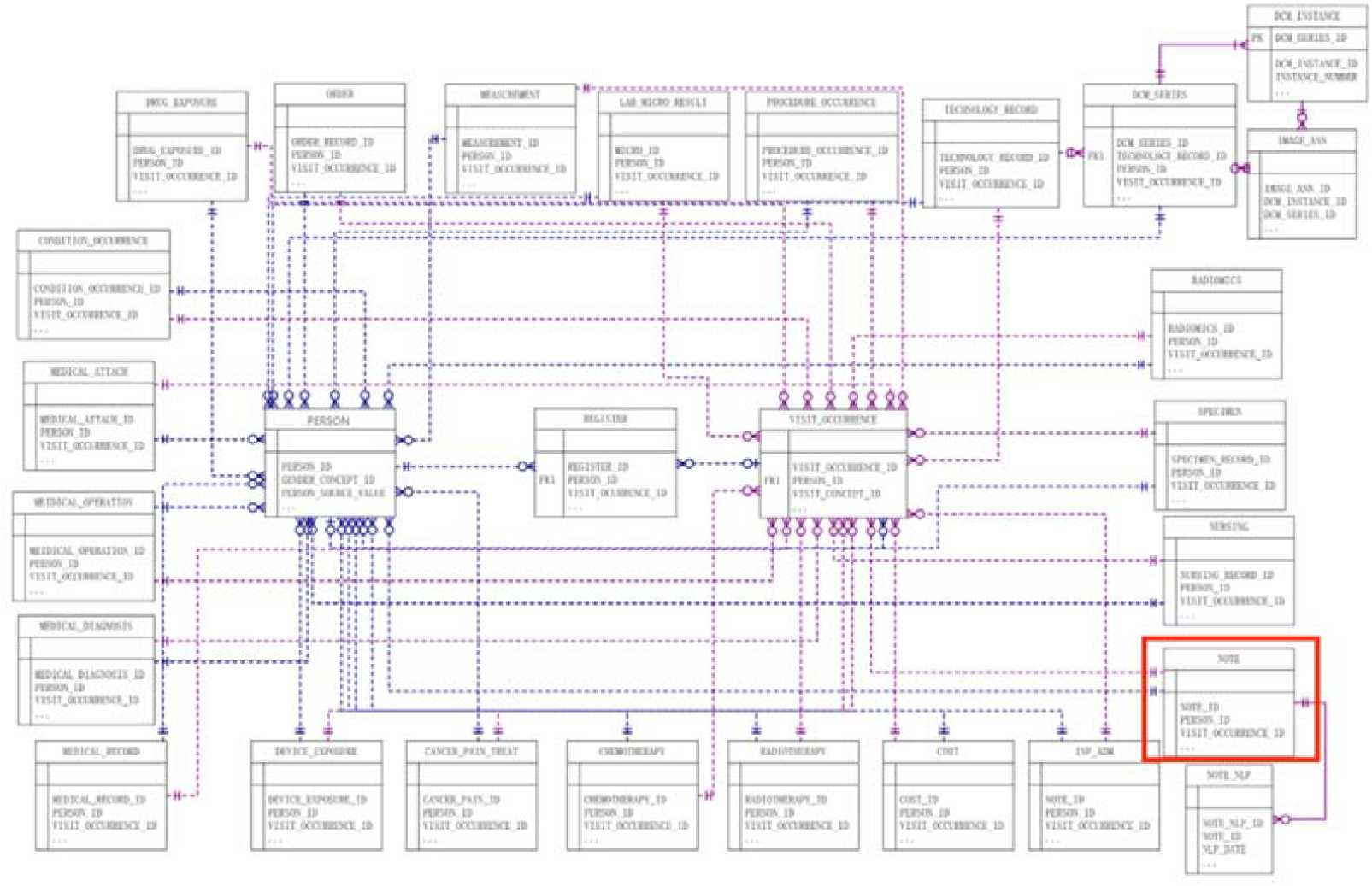
Structure of PUMCH’s disease-specific data platform. CDM structure suitable for Chinese EMRs under the OMOP system.

For structured data, field-specific anonymization is achieved through predefined table schemas. Consequently, sensitive information extraction and protection focus exclusively on unstructured textual fields within Chinese EMRs—primarily the aforementioned 13 documentation types stored in the NOTE master table.

From 4 million raw textual records, we randomly sampled 10,000 entries: 8,000 for training/validation and 2,000 for testing, enabling dual validation of information extraction accuracy and privacy strategy effectiveness.

### LLM Tools

The present study focuses on two primary objectives: first, identifying and marking patient-sensitive information within given long-form texts, and second, categorizing the identified sensitive information by risk severity levels. We have selected the Qwen2.5-72B-Instruct model [11] as our foundational architecture - a 72-billion-parameter open-source LLM developed by Alibaba Cloud. This model excels in deep comprehension of multi-step, cross-modal complex instructions, enhanced by ultra-long context processing and RLHF optimization techniques, making it particularly suitable for accurately parsing ambiguous requirements and generating structured outputs that align with our operational goals.The figure 2 below shows our research roadmap.

**Figure 2.**
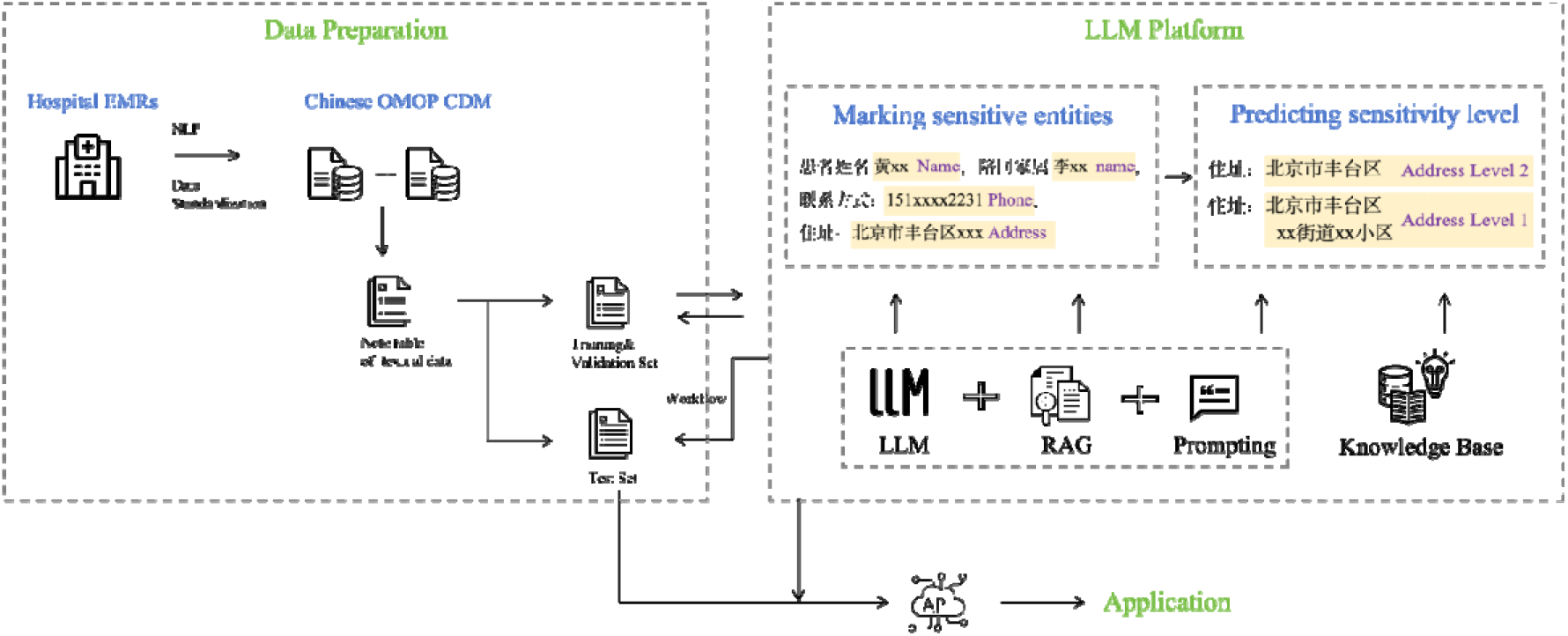
Research roadmap. The research roadmap illustrates three key steps: data preparation, large model processing and API application.

Notably, this research intentionally avoids engineering techniques like model fine-tuning or knowledge distillation, instead accomplishing its objectives solely through Prompt engineering and Retrieval-Augmented Generation (RAG). In LLM systems, Prompts serve as natural language instructions that guide model responses by decoding core human intent and contextual understanding. Their key strength lies in enabling flexible natural language interactions without requiring specialized formatting or programming syntax, thereby directly translating complex semantic content into contextually appropriate outputs. RAG enhances output authenticity and reliability through a two-phase process: first retrieving high-quality information from external knowledge bases, then generating answers based on verified data. This approach leverages dynamic retrieval mechanisms to anchor responses in up-to-date or domain-specific knowledge, effectively mitigating model hallucinations while improving result verifiability and accuracy.

Our Prompt implementation employs specially formatted natural language instructions, while RAG operations are facilitated through the open-source bisheng platform [12]. Bisheng provides a comprehensive LLM application development environment featuring modular tools for knowledge base management, agent orchestration, and skill reuse. Its visual interface streamlines model deployment workflows, with core capabilities centered on integrating RAG technology with dynamic knowledge repositories to significantly enhance response precision. The knowledge base employs the BGE-large-zh-v1.5 embedding model [13] from Beijing Academy of Artificial Intelligence, optimized specifically for Chinese semantic similarity calculations and retrieval tasks. Through bisheng’s visual interface, we combine natural language Prompts with our proprietary repository of sensitive information patterns to enable no-code LLM application development - the foundational approach for all subsequent research activities in this project.

### Marking of Sensitive Information

Our primary task is to leverage large language models to identify and label sensitive information within EMRs text entries. Based on our previous research, the desensitized information identified in this task is mainly divided into two major categories encompassing seven entity types. The first category pertains to basic privacy information, including names, national ID numbers, contact information, residential addresses, and healthcare facility names. This type of information is not distinguished between patients and medical staff, meaning that the relevant information of physicians will also be recognized. The second category relates to patients’ medical sensitive information, including pregnancy and childbirth-related information[14], as well as information related to STDs[15]. The labeling of sensitive information primarily relies on prompt engineering techniques.

According to the application domains, prompt engineering can be classified into twelve major categories[16]. For our task, we mainly utilize capabilities such as New Tasks Without Extensive Training, Reasoning and Logic, and Reduce Hallucination. For names, ID numbers, contact information, and addresses, we adopt the Few-shot Prompting approach[17]. For hospital names, pregnancy-related privacy information, and STD-related information, we employ RAG[18]. Finally, we use Chain-of-Thought (CoT) Prompting[19] to complete all entity labeling tasks in a single prompt.

We begin by individually labeling each entity category for sensitive information and iteratively refine the prompts based on results generated from real training data until the optimal performance is achieved on the validation set. Once optimal prompts for all seven entity types have been developed, we then merge them into a single prompt using CoT prompting to enable simultaneous labeling of all sensitive information within a text.

For the four entity types — names, ID numbers, contact information, and addresses — we adopt Few-shot Prompting. This technique guides the large model to accurately identify target entities by designing concise prompt templates and incorporating a few annotated examples. Chinese names, ID numbers, contact information, and addresses generally follow fixed patterns (e.g., 18-digit combinations for ID numbers, administrative divisions in addresses, and numerical patterns for phone numbers), enabling the large model to perform high-precision entity extraction with low complexity.

For the hospital name entity, we primarily utilize the RAG method. The core advantage of RAG lies in its knowledge augmentation and disambiguation capabilities. By retrieving external medical knowledge bases (e.g., hospital directories and standardized institutional codes) and combining them with model inference, it can accurately identify implicit variants of hospital names in text (such as abbreviations, aliases, or non-standard expressions), while also dynamically verifying the validity of the entities (e.g., matching with official registration information), effectively mitigating the issue of model hallucination. Additionally, the knowledge base can be updated in real time to accommodate newly established institutions, significantly improving the accuracy and reliability of entity extraction.

For pregnancy-related privacy information and STD-related information, we also employ the RAG approach. Our previous work has already accumulated partial corpora for these two categories. Furthermore, we can directly construct a knowledge base using common Chinese ICD-9 and ICD-10 disease descriptions. RAG separates the task into two modules: Retrieval and Generation. The retrieval module, using a lightweight model, quickly matches pregnancy and STD-related structured data in the knowledge base, while the generation module focuses on recognition and generation based on the retrieval results. This modular design allows for the independent optimization of each component, ensuring high-accuracy medical entity extraction while reducing computational load, making it more suitable for deployment in privacy-sensitive scenarios.

For all seven entity types, we define output formats within the prompts. The large model labels directly on the original text using ‘<<’ and ‘>>’ to mark positions, appending the corresponding entity type, and returns the labeled text without any unrelated output. If no sensitive information is detected, the model simply returns the original text. For example, “患者▪三……” would return “患者<<▪三: 姓名>>……”, while “患者性▪女……” would return “患者性▪女……”.

After achieving satisfactory extraction performance for all seven entity types on the training set using the individual prompts, we consolidate them into a single unified prompt using CoT prompting to enable simultaneous extraction. This unified prompt is further refined using the validation set to obtain optimal results. The final prompt is then evaluated on the test set by calculating precision and recall.

### Protection of Sensitive Information

Through the aforementioned steps, we have successfully extract sensitive information from the text. Given the importance of sensitive information, our priority is to maximize the recall rate of extraction in order to reduce omissions. However, this approach may affect the usability of the desensitized data, as it may result in the identification of data that is not particularly sensitive but is important for subsequent research purposes. Therefore, we further assess the extracted sensitive information to determine its sensitivity level, and then perform desensitization according to different sensitivity levels based on the data usage requirements of the subsequent research.

The assessment of sensitivity levels is not applied to all the extracted entity types. For the four types of entities—name, ID number, contact information, and hospital name—the sensitivity level is uniformly set to the highest level, and no further classification is required. For address entities, two levels are defined: Level 1 includes specific address information, such as street names, residential community names, or door numbers; Level 2 includes only information up to the district or county level, such as “Haidian District, Beijing.” For sexually transmitted disease (STD) information and pregnancy-related privacy information, three privacy levels are defined: Level 1 is highly sensitive, including information that could lead to serious social discrimination if disclosed, such as HIV/syphilis infection or miscarriage records; Level 2 is moderately sensitive, which may affect social perception if disclosed, such as mycoplasma/herpes infections or pregnancy-related surgical records; Level 3 is mildly sensitive, containing indirect descriptions of a patient’s condition, such as hepatitis B or C infections, or marital and childbearing status.

As seen from the definitions above, our classification of privacy levels adopts a qualitative approach with no clearly defined boundaries, which differs from traditional methods that rely on specific and explicit definitions. Here, we aim to leverage the large model’s inherent comprehension ability to perform this task through natural language descriptions, and evaluate the model’s performance in such tasks, which are common across various aspects of the healthcare domain.

For the sensitivity classification of location, STD information, and pregnancy-related privacy information, we follow the same methodology as the entity extraction task. That is, we first optimize the classification for each entity type on the training set, then combine the tasks and revise them through a second round on the validation set, and finally calculate the micro-average score on the test set.

## Results

### Use of LLM

All model development tasks in this study were carried out on the Bisheng platform and mainly included the following three types of tasks:

Knowledge Base Construction: We uploaded three types of dictionaries to the platform—hospital name lists, sexually transmitted disease lists, and pregnancy-related privacy description lists. The platform automatically parsed and segmented them to form structured knowledge bases.

Prompt Construction: For all large model tasks, prompts were designed and then uniformly categorized into two types of prompts.

Workflow Development: Two separate workflows were established, one for information tagging and the other for sensitive information classification. These workflows support end-to-end processing from raw text to final results. Additionally, the platform encapsulated the entire process into APIs, allowing real-world environments to directly access the processing via API calls.

### Sensitive Information Tagging Results

Table 1 illustrates the precision and recall performance for the seven types of entities on the test set. Compared with early neural network-based NER systems, achieving over 95% precision and over 90% recall across the seven entity types represents a substantial advancement. For instance, in earlier sequence labeling models, the LSTM-CNN method by Chiu and Nichols (2016) reported an F1 score of only 86.17% [20]. This result fully demonstrates the reliability of using large model techniques for entity recognition tasks.

**Table 1.** Precision and Recall for Tagging Seven Types of Entities on the Test Set.

| Label Class | Entity Class | Precision | Recall |
| --- | --- | --- | --- |
| Basic Information | Name | 98% | 98% |
|  | ID-code | 98% | 98% |
|  | Phone | 98% | 98% |
|  | Address | 98% | 98% |
|  | Hospital Name | 96% | 98% |
| Medically sensitive information | Pregnancy and Gestation Information | 95% | 90% |
|  | Sexually Transmitted Infections Information | 95% | 90% |

Notably, LLMs exhibit significantly superior performance on basic information entities than on medical information entities. Large language models are typically more adept at extracting basic entities such as names and addresses because their pretraining corpora (e.g., large-scale crawled web texts and books) contain a wealth of such examples. In contrast, specialized medical terms—such as those related to sensitive diseases—occur far less frequently, resulting in weaker representation capabilities and vocabulary gaps [21].

Furthermore, despite supplementary dictionaries via the RAG approach, this still cannot fully address the limitations in representational capacity. Additionally, medical entities often follow complex naming conventions, with abundant synonyms and abbreviations (e.g., HPV for human papillomavirus), and accurate disambiguation requires precise clinical context. General-domain LLMs, if not fine-tuned for specific fields, often misidentify or overgeneralize these terms [22].

### Prompt for Sensitive Information Tagging

To effectively identify sensitive entities—specifically hospital names, patient pregnancy-related privacy information, and patient sexually transmitted disease (STD) information—we constructed dedicated retrieval dictionaries for each category, which were used to build corresponding vector databases. Full dictionary entries are provided in Appendix 1.

The hospital name dictionary includes full names, abbreviations, and acronyms (in both Chinese and English) for the majority of hospitals across China.

The pregnancy-related privacy dictionary comprises descriptive phrases concerning obstetric history, pregnancy-related clinical notes, and fetal condition records.

The STD-related dictionary contains a list of sensitive diseases with both English and Chinese abbreviations.

For the complete set of seven entity types, we designed a unified Prompt leveraging a Chain-of-Thought (CoT) reasoning approach. This Prompt guides the model to label entities step-by-step based on their category within the original text and outputs the annotated result in a fixed return format, which ensures the consistency and standardization required for downstream processing.

We further enhanced model performance by using Few-shot Prompting, providing several clear in-context examples to help the large language model better understand the task and improve recognition accuracy. The complete prompt text can be found in Appendix 2.

Based on the above dictionaries and prompt structure, we utilized the Bisheng platform’s built-in workflow template to construct an automated sensitive data tagging pipeline. This pipeline supports export in JSON format and also offers a packaged API, enabling seamless integration with training and validation workflows. The complete data pipeline JSON file is included in Appendix 3.

### Sensitive Information Sensitivity Level Classification Results

After labeling the seven categories of entities, we performed a sensitivity level classification task based on the annotated text. The classification schema is defined as follows:

For names, ID numbers, contact information, and hospital names, the sensitivity level is uniformly set to Level 1 (Highly Sensitive). No further classification is required.

For addresses, we define two levels:

Level 1 (Highly Sensitive): Information that includes specific details such as street names, residential areas, or house numbers.

Level 2 (Moderately Sensitive): Information limited to county or district-level regions, such as “Haidian District, Beijing”.

For sexually transmitted disease (STD) information and pregnancy-related privacy information, we define three levels:

Level 1 (Highly Sensitive): Includes conditions that may result in severe social stigmatization if disclosed, such as HIV or syphilis, or extremely private records such as abortion history.

Level 2 (Moderately Sensitive): Includes conditions that may affect social perception, such as mycoplasma or herpes, or procedural records related to childbirth.

Level 3 (Mildly Sensitive): Indirect or less stigmatizing health information, such as hepatitis B/C or marital and reproductive status.

As this is a multi-class classification task, we used the micro-average score to evaluate model performance in terms of both precision and recall. Given that sensitivity level classification lacks clear quantitative thresholds, correctness is primarily determined by expert judgment. To enhance annotation reliability, we adopted a dual-expert cross-validation approach:

If the two experts agreed on a classification, that result was taken as the ground truth.

In cases of disagreement, the lower sensitivity level was chosen as the final label, in order to support broader data usability and avoid over-sanitization.

Model predictions were then compared with these ground truth labels to calculate classification performance. Table 2 presents the micro-average precision and recall for the three entity types subject to sensitivity level classification.

**Table 2.**
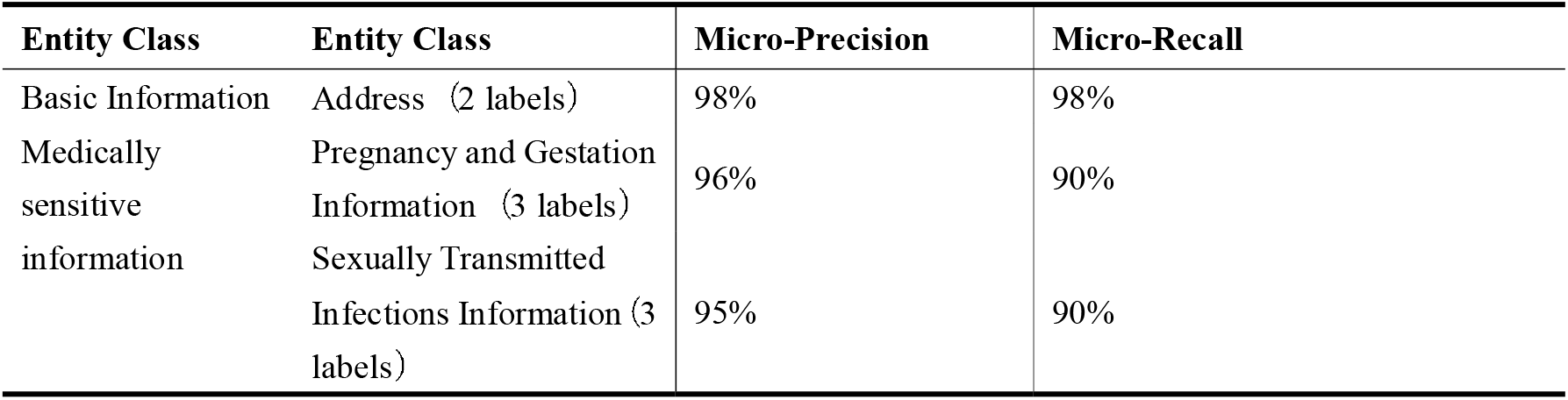
Micro-average Precision and Recall for Sensitivity Level Classification of Three Entity Types on the Test Set.

### Prompt for Sensitive Information Sensitivity Level Classification

For the task of sensitivity level classification, we adopted a Chain-of-Thought (CoT) prompting approach. This method processes the previously labeled text and determines the sensitivity level for each entity. The sensitivity level is appended directly after the entity label in the annotated text:

Names, ID numbers, contact information, and hospital names are uniformly labeled as Level 1 (Highly Sensitive).

The remaining entity types—addresses, STD-related information, and pregnancy-related privacy information—are classified by the model according to predefined criteria.

The complete prompt used for this task can be found in Appendix File 2.

Based on this prompt, we developed a sensitivity level classification workflow for private data. The full data flow in JSON format is available in Appendix File 3, and the workflow also supports direct API invocation, allowing for streamlined integration into real-world systems.

By combining the workflows for sensitive information tagging and sensitivity level classification, we enable a fully automated pipeline for sensitive data protection. This setup supports targeted data de-identification based on the required sensitivity level for subsequent research use, allowing researchers to balance privacy and data utility as needed.

## Discussion

### Principal Findings

In this study, we have innovatively developed a no-code privacy protection solution based on large language models (LLMs), utilizing natural language interactions to intelligently de-identify medical data. The solution incorporates a three-tier protection mechanism that dynamically adapts privacy preservation strategies to clinical scenario demands, maximizing data value retention while ensuring data security. This approach provides important insights for applying LLMs in medical data governance and showcases an innovative path for combining AI technology with medical compliance requirements.

Breakthrough Achievements: This research marks a breakthrough in applying large language models in the field of sensitive information protection, particularly within medical research contexts. Compared to traditional AI models that require specialized algorithm engineers, large language models can handle complex data de-identification tasks through natural language interactions. This significantly lowers the AI adoption threshold for healthcare institutions.

Unique Value of Data De-identification: When it comes to privacy protection techniques, data de-identification presents distinct practical value. Compared to the technical complexity of federated learning or the high computational costs associated with data encryption, large model-based intelligent de-identification systems maintain the statistical analysis value of the original data. Additionally, these systems leverage natural language processing capabilities to provide context-sensitive and precise masking of sensitive information within medical records.

We have used large model technology to address traditional entity recognition and classification tasks, achieving good results overall. This success encourages us to explore other types of tasks in the future. As large models continue to evolve, embracing these technologies to better serve our research remains a substantial challenge for healthcare professionals.

### Limitations

Despite the promising outcomes of this study, there are still several limitations that need to be addressed:

Data Limitations: The model’s test and validation datasets were sourced excusively from Beijing Union Medical College Hospital. Although we strictly divided the training, validation, and test sets, the singular data source may limit the model’s generalization ability. Future research should include data from multiple healthcare institutions (e.g., top-tier hospitals, primary care hospitals) and perform multi-center validation to comprehensively assess the model’s practical value.

Lack of Privacy Rule Adjustment Mechanisms: The current privacy protection scheme relies mainly on static rules defined by sensitivity level parameters, lacking the capability for dynamic adjustments. Additionally, the instability of large model outputs may trigger systemic risks—any changes in the rules can have cascading effects, requiring a reevaluation of the entire system’s security. This necessitates the development of more stable and adaptive privacy protection frameworks.

De-identification Precision Issues: Existing de-identification algorithms suffer from both over-de-identification and under-de-identification problems. Over-de-identification can lead to the loss of crucial clinical information, severely affecting data usability, while under-de-identification may result in sensitive information being overlooked. More accurate semantic recognition technology is needed to protect privacy while maximizing data value retention. It is recommended to introduce manual review mechanisms as a transitional solution.

### Future Directions

The application of large language models in healthcare has already permeated multiple stages of the clinical workflow, ranging from fundamental data cleaning and standardization to complex clinical decision-support systems. The breadth and depth of these applications continue to expand. However, this technological penetration presents dual challenges: on one hand, the accuracy and stability of model outputs directly affect the quality of medical services; on the other hand, the unique nature of medical scenarios demands that algorithms be highly reliable and interpretable. In future research, we will focus on the following:

Application Expansion: Exploring the potential applications of large models in emerging areas such as personalized treatment plan generation, assistive diagnostics, and healthcare resource optimization.

Reliability Validation Framework: Establishing a comprehensive evaluation framework that includes clinical effectiveness, algorithm stability, and ethical compliance.

Human-AI Collaboration Mechanism: Investigating how to combine the predictive power of large models with clinical experts’ experience and judgment to build more intelligent decision support systems.

Through continuous technological iterations and rigorous clinical validation, we aim to transform large models into truly reliable research tools—leveraging their strengths in data processing and pattern recognition while ensuring their outputs meet the high standards required in medical practice. The deep integration of this technology with medicine will open new possibilities for enhancing healthcare quality and fostering research innovation.

## Conclusions

We have innovatively developed a no-code privacy protection solution based on large language models, which enables intelligent de-identification of medical data through natural language interaction. This solution adopts a three-level hierarchical protection mechanism, which can automatically match the appropriate privacy protection strategies according to clinical scenario requirements, ensuring data security while maximizing the retention of the data’s research and application value.

This practice provides an important reference for the application of large models in medical data governance and demonstrates an innovative path for the integration of AI technology with medical compliance requirements. It not only provides the medical industry with a more convenient privacy protection method but also promotes the further development and application of large language models in the medical field. Through this technology, we can achieve efficient de-identification of data, providing more reliable protection for the secure sharing of medical data and clinical research.

## Data Availability

The datasets supporting the findings of this study are available from the corresponding author upon reasonable request. The hospital data cannot be made publicly available due to its sensitive nature.

